# Unraveling the Efficiency of Non-Navigated 2D Intraoperative Ultrasound in Glioma Surgery: Challenging the Demand for Increased Technological Sophistication in Intraoperative Imaging

**DOI:** 10.1101/2023.06.01.23290675

**Authors:** Santiago Cepeda, Sergio García-García, Ignacio Arrese, Rosario Sarabia

## Abstract

**Background:** In an era characterized by rapid progression in neurosurgical technologies, traditional tools such as the non-navigated two-dimensional intraoperative ultrasound (nn-2D-IOUS) risk being overshadowed. Against this backdrop, this study endeavors to provide a comprehensive and rigorous assessment of the clinical efficacy and surgical relevance of nn-2D-IOUS, specifically in the context of glioma resections.

**Methods:** This retrospective study undertaken at a single center evaluated 99 consecutive, non-selected patients diagnosed with both high-grade and low-grade gliomas. The primary objective was to assess the proficiency of nn-2D-IOUS in generating satisfactory image quality, identifying residual tumor tissue, and its influence on the extent of resection. To validate these results, early postoperative MRI data served as the reference standard.

**Results:** The nn-2D-IOUS exhibited a high level of effectiveness, successfully generating good quality images in 79% of the cases evaluated. With a sensitivity rate of 68% and a perfect specificity of 100%, nn-2D-IOUS unequivocally demonstrated its utility in intraoperative tumor detection. Notably, in cases where total tumor removal was the surgical objective, a resection exceeding 95% of the initial tumor volume was achieved in 86% of patients. Additionally, in cases where residual tumor was not detected by nn-2D-IOUS, the mean volume of undetected tumor tissue was remarkably minimal, averaging at 0.29 cm^3^.

**Conclusion:** Our study provides compelling evidence supporting the invaluable role and efficacy of nn-2D-IOUS in glioma surgery. The results underscore the potential of harnessing traditional, cost-effective technologies such as nn-2D-IOUS to achieve enhanced surgical outcomes, even in the face of more advanced alternatives. These insights carry significant implications, particularly for resource-constrained settings, emphasizing the importance of optimizing the use of existing tools to improve patient care in a practical and efficient manner.

## INTRODUCTION

Gliomas, the most prevalent primary brain tumors, account for approximately 30% of all central nervous system neoplasms and 80% of all malignant brain tumors ^1^. The extent of resection (EOR) significantly influences the outcome of glioma surgery, with studies demonstrating a direct correlation between EOR and overall survival ^2^. However, achieving maximal safe resection is often challenging due to the infiltrative nature of gliomas and their proximity to eloquent brain areas. As a result, various intraoperative adjuncts have been developed to aid neurosurgeons in this task.

Intraoperative ultrasound (IOUS) has been instrumental in providing real-time imaging, allowing for improved visualization of tumor margins during surgery ^3–5^. Its affordability, and portability, render it a valuable tool in the neurosurgical armamentarium. Despite its merits, non-navigated two-dimensional intraoperative ultrasound (nn-2D-IOUS) has faced criticism due to its steep learning curve and its reliance on the surgeon’s ability to mentally reconstruct a three-dimensional image from the two-dimensional representation. Intraoperative MRI (iMRI), another imaging tool, has also been utilized to enhance visualization, although its application is accompanied by a significant increase in costs and logistical complexity ^6,7^.

In recent years, the landscape of neurosurgical technology has been marked by a rapid evolution towards more advanced, and often more expensive, modalities. This transition, fostered by industry influence and the drive for innovation, has introduced three-dimensional (3D) and navigated IOUS systems, among others, to the neurosurgical theatre ^8–10^. These advancements, while addressing several challenges presented by conventional imaging modalities, have inadvertently overshadowed the perceived utility of the nn-2D-IOUS, leading to a substantial decrease in its use. Given these considerations, the focus of this investigation is to critically evaluate the efficacy and utility of nn-2D-IOUS in glioma surgery. Through a meticulous analysis of a consecutive case series, we aim to elucidate the impact of this traditional modality on the extent of resection and subsequent patient outcomes. This inquiry is not only a retrospective appraisal of nn-2D-IOUS but also an invitation to the scientific community to reassess the need for high-cost, technologically advanced systems. As we strive for maximum safe resection, the question arises: could the more accessible and cost-effective nn-2D-IOUS be sufficient? Our aim is to refocus attention on this technology and instigate a data-informed discussion regarding the ideal equilibrium between technological sophistication and clinical efficacy within the field of glioma surgery.

## METHODS

### Study population

A retrospective analysis was conducted on consecutively and non-selected operated patients between June 2018 and December 2022 in our neurosurgical department, with a confirmed histopathological diagnosis of glioma (including high and low-grade gliomas). The inclusion criteria were as follows: adult patients over 18 years of age who underwent craniotomy, intraoperative ultrasound assessment, pre-operative magnetic resonance imaging (MRI), and early postoperative MRI (within 72 hours). Stereotactic biopsies and patients without postoperative MRI evaluation of the EOR were excluded. Demographic, clinical, treatment, and follow-up variables were collected using standardized methods. This study adhered to the Strengthening the Reporting of Observational Studies in Epidemiology (STROBE) guidelines, obtained approval from the local ethics committee (Ref. 21-PI085), and was registered on ClinicalTrials.gov, Identifier: NCT05873946.

### Intraoperative acquisition technique

All cases underwent craniotomy with careful consideration of its dimensions to ensure proper placement and maneuverability of the ultrasound probe. The initial ultrasound image acquisition took place after craniotomy but before dural opening. We utilized a Hitachi Noblus ultrasound system with a C42 micro convex probe operating within a frequency range of 4 to 8 MHz. The scan width was set at a 20 mm radius, and the field of view scan angle was 80°. To maintain sterility, the probe was covered with double sterile sheets, and a minimal amount of conductive gel was applied within the sheet.

Systematic acquisition was performed initially in B-mode, capturing images in various orthogonal planes depending on the tumor location and identifying anatomical structures that could serve as references. Subsequently, the microvascular Doppler mode was used to assess the tumor’s vascularity and its relationship with arterial branches that could serve as landmarks. Finally, elastograms were acquired to evaluate the tumor’s consistency using the previously described technique ^11^. All intraoperative ultrasound studies were conducted by the same surgeons (S.C and R.S). The number of acquisitions necessary for each case was determined by the surgeon, with a minimum of two studies performed: one before tumor resection and another after resection. Additional acquisitions could be performed as deemed necessary during the tumor resection process. An illustrative case is shown in Figure 1.

**Figure 1.**
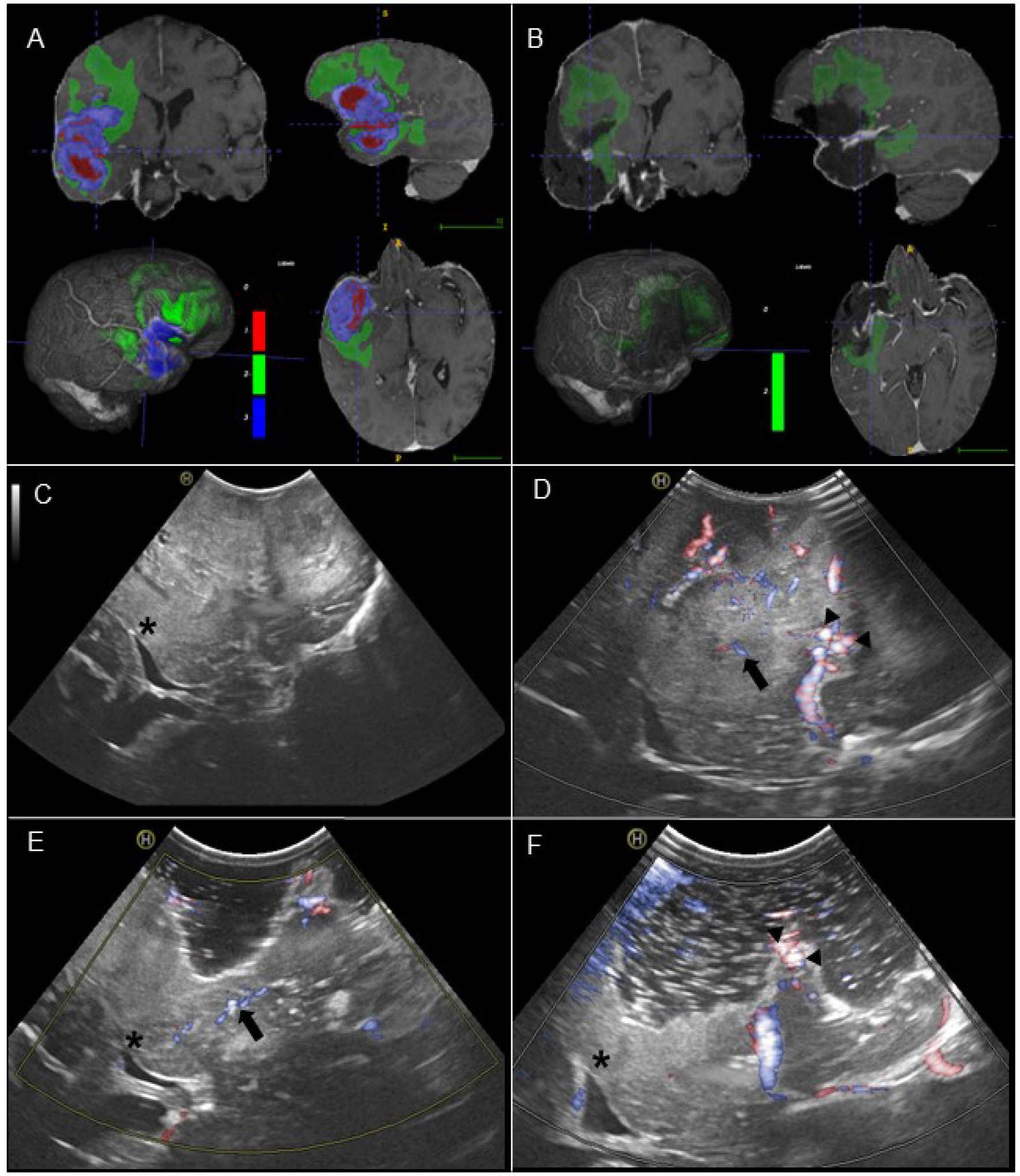
An illustrative of a patient with a peri-sylvian right wild-type grade 4 astrocytoma. The figure displays the results of preprocessing and automatic segmentation of tumor subregions on preoperative (A) and postoperative (B) MRI scans. The segmented regions are color-coded, with red representing necrosis, green indicating the peritumoral region, and blue denoting the enhancing tumor. Additionally, intraoperative ultrasound images (C and D) captured at the beginning of the surgery are presented. Image C is displayed in B mode, while image D utilizes an ultrasensitive Doppler mode. The ultrasound images highlight important anatomical landmarks, including the lateral ventricle (black asterisk), lenticulostriate arteries (black arrow), and M2 branches of the cerebral medial artery (CMA) (black head arrows). Images E and D were acquired at the end of the resection, revealing no residual tumor and improved patency of the perforating arteries following relief of mass effect. Furthermore, the distal branches of CMA demonstrate enhanced permeability, accompanied by a large surgical cavity surrounding the vessels, as confirmed by the postoperative MRI images.

### Assessment of intraoperative ultrasound images

The quality of intraoperative ultrasound images was evaluated and recorded using a subjective scale ranging from “Good quality,” “Medium,” to “Low quality.” Additionally, the presence of tumor borders was determined using the “Good,” “Medium,” and “Poorly defined” scale. The number of ultrasound examinations performed, and the presence or absence of residual tumor identified using this technique were also recorded.

### MRI scans processing and evaluation of the extent of resection

Pre- and postoperative MRI images were procured from the Picture Archiving Communication System (PACS) in the Digital Imaging and Communications in Medicine (DICOM) format, which were then processed. As an initial step, these images underwent conversion into the Neuroimaging Informatics Technology Initiative (NIfTI) format. Subsequently, volumetric measurements of the initial tumor size were executed, defined as the contrast-enhancing tumor for high-grade gliomas and the T2/FLAIR abnormality for low-grade gliomas. In addition, volumetric assessments of the peritumoral region, inclusive of both edema and infiltration, were integrated for glioblastomas.

Such measurements were further carried out in the postoperative study to determine residual volume and ascertain the extent of resection, which were categorized as follows ^12,13^:

- Gross total resection (GTR): Complete resection of the contrast-enhancing tumor for glioblastomas and total removal of T2/FLAIR hyperintensity for grade 2 and 3 gliomas.
- Near-total resection (NTR): Over 95% resection of contrast-enhancing tumor with less than 1 cm^3^ of residual tumor for glioblastomas or above 90% removal of T2/FLAIR tumor hyperintensity with less than 5 cm^3^ remaining for grade 2 and 3 gliomas.
- Subtotal resection (SR): Above 80% removal of contrast-enhancing tumor with less than 5 cm^3^ of residual tumor for glioblastomas and more than 40% reduction of T2/FLAIR hyperintensity with less than 25 cm^3^ left for grade 2 and 3 gliomas.
- Partial resection (PR): Between 1-79% resection of contrast-enhancing tumor with more than 5 cm^3^ of residual tumor for glioblastomas and between 1-39% reduction of T2/FLAIR hyperintensity with over 25 cm3 remaining for grade 2 and 3 gliomas.

Segmentation tasks were carried out automatically employing DeepBraTumIA (https://www.nitrc.org/projects/deepbratumia/), with the generated segmentations meticulously refined by experienced neurosurgeons, S.C. and S.G.

### Tumor Location Analysis: Mapping the Spatial Distribution and Estimating the Resectability Index

The Radionics software toolkit ^14^ was utilized as the cornerstone for conducting tumor location analysis and gauging the potential for resectability for the glioblastoma subgroup of patients. This method underwent a series of preprocessing stages. In the initial stages, resampling was carried out to ensure an isotropic spacing of 1 mm^3^. This step was succeeded by image refinement, either through meticulous trimming around the patient’s head to eliminate irrelevant background, or via skull-stripping using a custom-made brain segmentation model. The volume was subsequently re-adjusted to 128 × 128 × 144 voxels, aided by first-order spline interpolation. Concluding the process, intensity normalization was executed, confining the range between 0 and 1.

The process of standardizing preoperative clinical reports was systematically reproducible, involving the computation of tumor characteristics post alignment to a universally recognized reference space. This reference space was designated by the symmetric Montreal Neurological Institute ICBM2009a atlas (MNI) ^15^.

For each tumor, an extensive array of features was gathered, encompassing volume, laterality, multifocality, and locational profiles of cortical and subcortical structures. Utilizing these spatiotemporal features as critical parameters, an estimated resectability index was subsequently calculated, providing valuable insight into prospective surgical interventions.

### Surgical technique and operative adjuncts

All patients underwent surgery employing a microsurgical approach complemented by intraoperative ultrasound. The choice of intraoperative tools was tailored according to the surgeon’s (S.C.) professional discretion and the distinct location of the tumor. A variety of auxiliary surgical adjuncts were documented, each utilized based on the unique demands of the case at hand. These adjuncts encompassed neuronavigation, intraoperative fluorescence (utilizing agents such as sodium fluorescein or 5-aminolevulinic acid), direct electrical stimulation in awake surgeries, and intraoperative neurophysiological monitoring.

### Assessing Postoperative Neurological Deficits and Survival Prognosis

If neurological deficits were observed during the immediate post-surgical neurological examination, they were regarded as novel deficits. A deficit that resolved within 30 days following the surgery was categorized as transient. A deficit was classified as permanent if it persisted beyond the 30-day follow-up period. The timeframe encompassing the initial 30 days following the surgical procedure was defined as the postoperative period.

Glioblastoma patients from the subgroup who were treated according to the Stupp protocol ^16^ and had at least one year of follow-up were used for the survival analysis. In the context of this study, the terms ‘Overall Survival’ (OS) and ‘Progression-Free Survival’ (PFS) were used to measure patient outcomes. Overall Survival was defined as the time from the date of diagnosis to the date of death. Alternatively, in cases where patients were alive at the end of the study period, the date of last follow-up was used. Progression-Free Survival, on the other hand, was described as the time from the diagnosis to the point of tumor progression. For patients who didn’t show signs of disease progression during the study period, the date of last follow-up was employed.

### Statistical Analysis

All statistical analyses were conducted using R version 4.0.5 (R Foundation for Statistical Computing). The capacity of intraoperative ultrasound for the detection of residual tumor was determined using early postoperative MRI as a reference. This allowed for the computation of sensitivity, specificity, negative predictive value, and positive predictive value. Survival analysis was carried out using stepwise Cox proportional hazards regression. Predictors for GTR were analyzed using multivariate logistic regression.

## RESULTS

### Patient demographics and tumor features

During the study period, 199 brain tumors were treated, of which 99 were identified as gliomas. The average age of the patients was 59.73 ± 12.39 years. The gender distribution was 62% male and 38% female. A significant majority of surgeries (86%) were planned with the intent of complete resection, while 5% aimed at debulking the tumor, and 9% were scheduled as open biopsies. The patient cohort included 18 low-grade gliomas (LGG) and 81 high-grade gliomas (HGG). The mean preoperative tumor volume was 34.94 cm^3^. A summary of the patients’ clinical characteristics and the findings from pre and postoperative MRI studies can be found in Table 1.

**Table 1.** Demographic data and tumor characteristics.

| Table 1. Demographic data and tumor characteristics |  |
| --- | --- |
| Variable | Value |
| Age | 59.7 ± 12.4 |
| Sex |  |
| Female | 37 (37.4%) |
| Male | 62 (62.6%) |
| Preoperative KPS |  |
| 60 | 11 (11.1%) |
| 70 | 26 (26.3%) |
| 80 | 36 (36.4%) |
| 90 | 25 (25.3%) |
| 100 | 1 (1%) |
| Previous treatment |  |
| No | 81 (81.8%) |
| Surgery | 7 (7.1%) |
| Surgery + CTX + RT | 11 (11.1%) |
| Aim of surgery |  |
| Biopsy | 9 (9.1%) |
| Debulking | 5 (5.1%) |
| Gross total resection | 85 (85.8%) |
| WHO grade |  |
| 2 | 18 (18.2%) |
| 3 | 23 (23.2%) |
| 4 | 58 (58.6%) |
| IDH status |  |
| Mutant | 43 (43.9%) |
| Wild-type | 55 (56.1%) |
| Tumor location (main lobe) |  |
| Frontal | 38 (38.4%) |
| Insular | 6 (6.1%) |
| Occipital | 9 (9.1%) |
| Parietal | 10 (10.1%) |
| Temporal | 33 (33.3%) |
| Cerebellar | 3 (3%) |
| Side |  |
| Left | 50 (51.5%) |
| Right | 49 (49.5%) |
| Eloquent |  |
| No | 59 (59.6%) |
| Yes | 40 (40.4%) |
| Multifocal |  |
| No | 79 (79.8%) |
| Yes | 20 (20.2%) |
| Tumor depth |  |
| < 2 cm | 67 (67.7%) |
| > 2 cm | 32 (32.3%) |
| US image quality |  |
| Good | 78 (78.8%) |
| Medium | 15 (15.2%) |
| Poor | 6 (6%) |
| Tumor borders identified by US |  |
| Good | 33 (33.3%) |
| Medium | 31 (31.3%) |
| Poor | 55 (55.4%) |
| Postoperative KPS |  |
| 50 | 4 (4.1%) |
| 60 | 7 (7.2%) |
| 70 | 16 (16.5%) |
| 80 | 33 (34%) |
| 90 | 36 (37.1%) |
| 100 | 1 (1.1%) |
| Postoperative neurological deficit |  |
| No | 62 (63.9%) |
| Transient | 19 (19.6%) |
| Minor persistent | 11 (11.3%) |
| Major persistent | 5 (5.2%) |
| Postoperative complication |  |
| No | 83 (83.8%) |
| Hematoma | 6 (6.1%) |
| Infarction | 2 (2%) |
| Infection | 7 (7.1%) |
| Other | 1 (1%) |
| Values are expressed in mean, standard deviation (SD), and percentages. KPS = Karnofsky Performance Score. WHO = World Health Organization. CXT = Chemotherapy. RT = Radiotherapy. IDH = Isocitrate Dehydrogenase. US = ultrasound |  |

**Table 2.** Volumetric analysis and extent of resection.

| **Table 2. Volumetric analysis and extent of resection** |  |  |  |  |  |
| --- | --- | --- | --- | --- | --- |
|  | **All operations**<br>**n = 99 (100%)** | **Planned GTR**<br>**n = 85 (85.9%)** | **WHO grade 2**<br>**n = 18 (18.2%)** | **WHO grade 3**<br>**n = 23 (23.2%)** | **WHO grade 4**<br>**n = 58 (58.6%)** |
| **Preoperative tumor volume (cm<sup>3</sup>)** | 39.9 ± 28.7 | 33.6 ± 27.2 | 30.2 ± 16.7 | 29.9 ± 30.7 | 38.4 ± 30.6 |
| **Preoperative T2/FLAIR peritumoral volume (cm<sup>3</sup>)** | 60.5 ± 43.7 | 63.2 ± 44.2 | - | - | 63.4 ± 43.8 |
| **Residual tumor volume (cm<sup>3</sup>)** | 1.79 ± 5.7 | 0.74 ± 2.1 | 3.8 ± 6.8 | 1.5 ± 2.9 | 1.2 ± 6 |
| **Residual T2/FLAIR peritumoral volume (cm<sup>3</sup>)** | 35.9 ± 28.6 | 35.5 ± 28.2 | - | - | 40.2 ± 28.6 |
| **Mean EOR** | 95.7 ± 10.9 | 97.4 ± 6.9 | 89.6 ± 18.7 | 93.9 ± 10.6 | 98.2 ± 6 |
| **EOR (categories)** |  |  |  |  |  |
| **GTR** | 59 (59.6%) | 59 (69.4%) | 8 (44.4%) | 10 (43.5%) | 41 (70.7%) |
| **NTR** | 14 (14.1%) | 14 (16.5%) | 4 (22.2%) | 3 (13%) | 7 (12.1%) |
| **STR** | 13 (13.1%) | 9 (10.6%) | 5 (27.8%) | 5 (21.7%) | 3 (5.2%) |
| **PR** | 4 (4.1%) | 3 (3.5%) | - | 2 (8.7%) | 2 (3.5%) |
| **Biopsy** | 9 (9.1%) | - | 1 (5.6%) | 3 (13.1%) | 5 (8.5%) |
| Values are expressed in mean, standard deviation (SD), and percentages. EOR = Extent of resection. WHO = World Health Organization. GTR = Gross total resection. NTR = Near total resection. STR = Subtotal resection. PR = Partial resection. |  |  |  |  |  |

### Extent of resection

From the total patient cohort, GTR was achieved in 59 patients (60%), NTR in 14 patients (14%), STR in 13 patients (13%), PR in 4 patients (4%), and biopsy only in 9 patients. The mean residual volume was 1.79 cm^3^. Within the subgroup of patients for whom a GTR was planned, it was successfully achieved in 69% of cases. A resection greater than 95% of the initial tumor volume (GTR + NTR) was accomplished in 86% of patients from this subgroup. In LGG, the achieved GTR rate was 44% for the total cohort and 53% in cases with radical surgery intent. For HGG, the GTR rate was 63% for the entire cohort and 73% for cases of planned total resection. For glioblastomas, the total GTR was 70% and 81% in resections with radical intent, which, when combined with NTR, reached 96%. The most frequently employed surgical support tools were neuronavigation (utilized in all but one case) and fluorescence with 5’ALA. (Figure 2).

**Figure 2.**
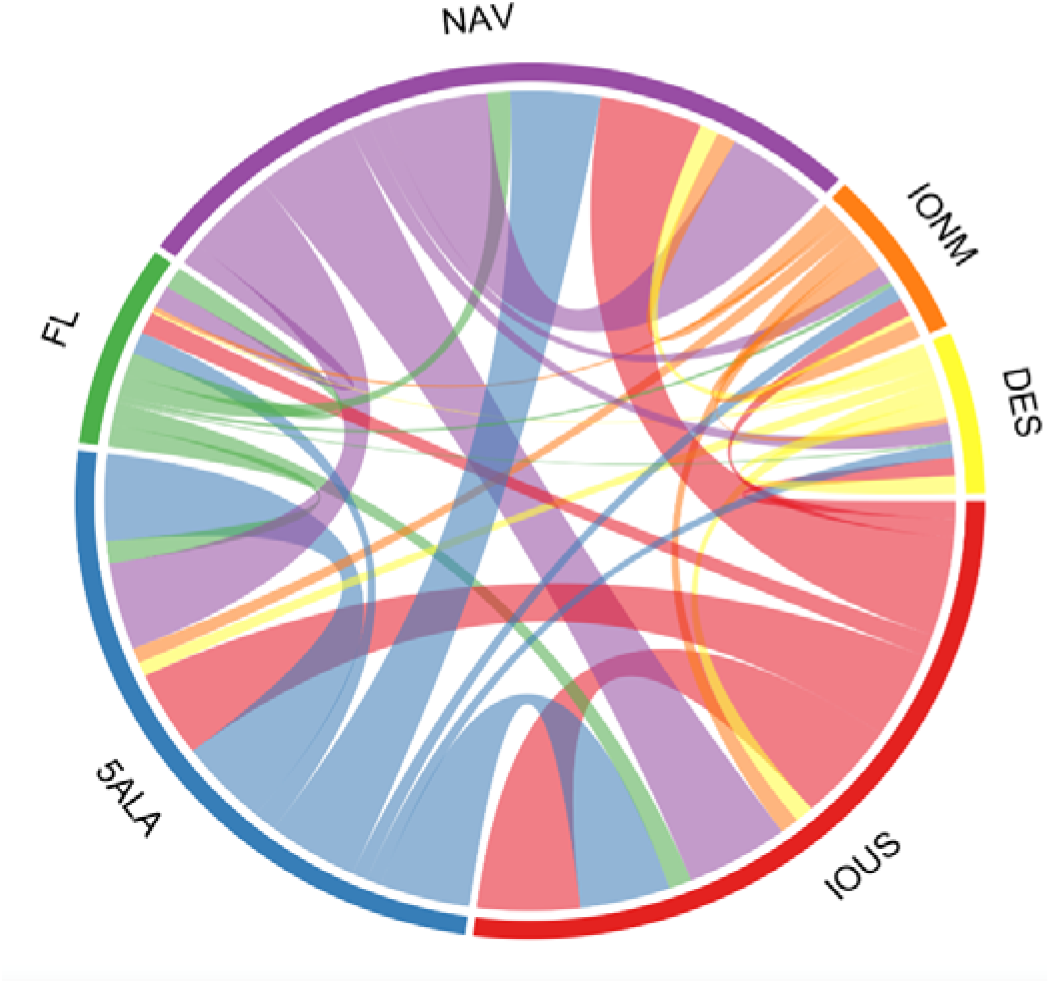
Chord diagram depicting the intricate interplay and synergistic utilization of operative adjuncts in the study. The diagram visually represents the combinations and relationships among different surgical adjuncts employed, providing a comprehensive view of their interconnected roles and contributions.

The multivariate logistic regression analysis identified tumors located in eloquent areas as the sole significant predictor for GTR exhibiting an odds ratio (OR) of 0.30 (95% CI [0.13, 0.69], p = 0.005). This result highlights that tumors situated in eloquent areas were significantly associated with a decreased likelihood of achieving GTR.

### Intraoperative Ultrasound Image Assessment

The average number of intraoperative ultrasound examinations performed was 2, ranging between 2 and 5. Image quality was categorized as good for 79% of the examinations, moderate for 15%, and poor for 6%. The identification of tumor borders was rated as good in 33% of cases, average in 31%, and poor in 35%. In 27% of cases, intraoperative ultrasound was able to discern residual tumor presence, while in the remaining 72% of cases, no residual tumor was detected using this technique.

### Neurological Impairments and Postsurgical Complications

Out of the total patient cohort, three deaths occurred due to postoperative complications. Transient neurological deficits were experienced by 19% of patients, while 11% suffered mild permanent deficits, and 5% exhibited severe permanent postoperative deficits. In addition, there were 7 recorded postoperative infections, 6 postoperative hematomas that did not require reoperation but extended the hospital stay, and 2 postoperative infarcts with clinical involvement.

### Comparison of Intraoperative Ultrasound with Early Postoperative MRI

In the early postoperative period, residual tumor presence was detected by MRI in 40% of patients. The effectiveness of intraoperative ultrasound (IOUS) in identifying residual tumor was evaluated using a confusion matrix, which demonstrated a sensitivity rate of 68%, a specificity of 100%, and a diagnostic accuracy of 87%. Moreover, the negative predictive value was determined to be 82%, while the positive predictive value achieved a perfect score of 100%. The inter-method agreement, measured by the Kappa value, yielded a value of 0.71. Additionally, the mean volume of residual tumor undetected by ioUS was remarkably minimal, measuring just 0.29 cm^3^. The results are summarized in Figure 3 and Table 3.

**Table 3.** Diagnostic performance of IOUS to detect residual tumor.

| Table 3. Diagnostic performance of IOUS to detect residual tumor |  |  |
| --- | --- | --- |
| Metrics | Mean | 95 % CI |
| Sensitivity | 67.5% | 57.5 – 76.7% |
| Specificity | 100% | 96.3 – 100% |
| PPV | 100% | 96.3 – 100% |
| NPV | 81.9% | 72.8 – 88.9% |
| Accuracy | 86.9% | 78.6% - 92.8% |
| Values are expressed in number of patients, percentages and 95% confidence interval. PPV = Positive predicted value. NPV = Negative predicted value. |  |  |

**Figure 3.**
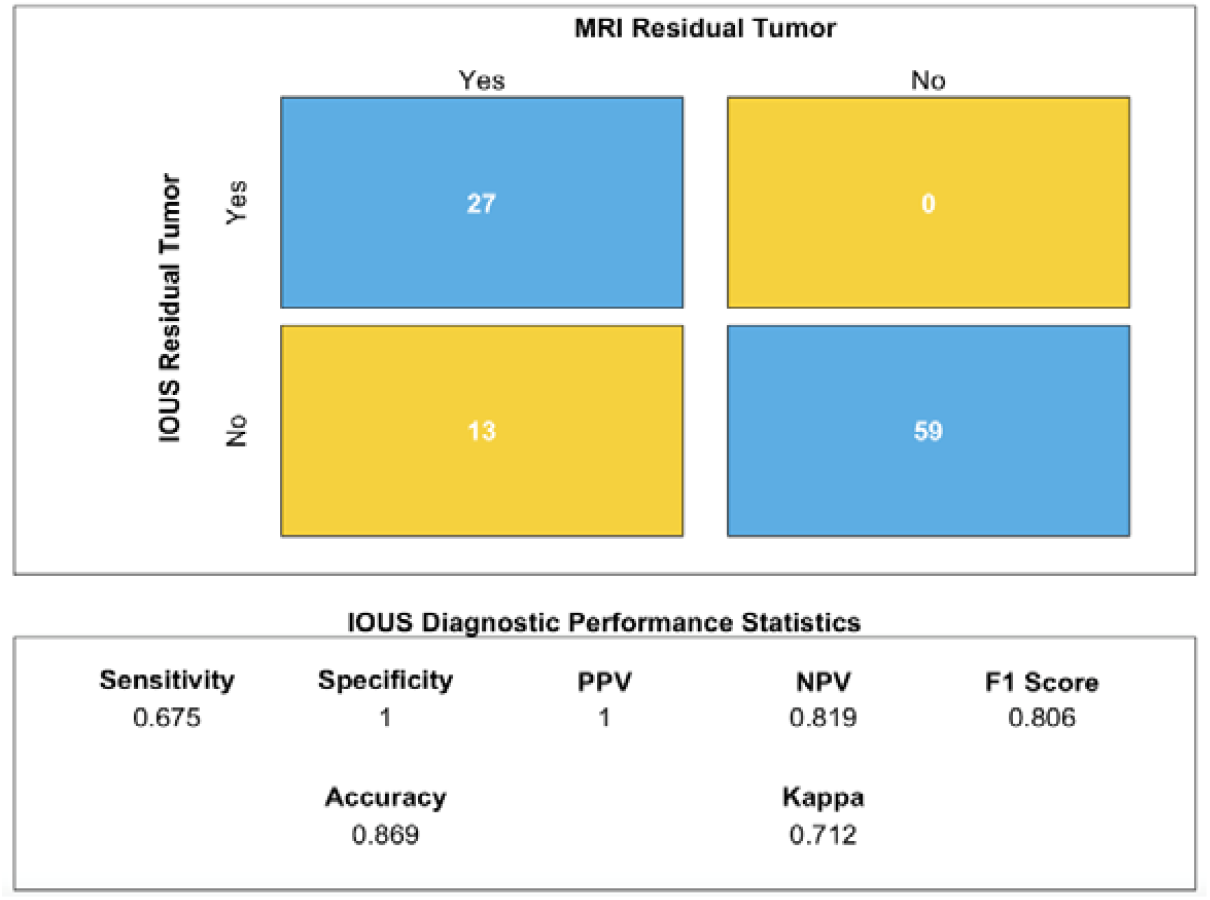
Confusion matrix illustrating the instances of residual tumor detected by intraoperative ultrasound (IOUS) compared to the reference image modality, early postoperative MRI. The matrix provides a comprehensive overview of the classification results.

### Estimating the Probability of Resectability in Glioblastomas

In our study, we utilized probabilistic maps provided by Raidionics to estimate the volume of residual tumor in a specific subgroup of glioblastoma patients. The results revealed an estimated residual volume of 7.20 ± 5.89 cm^3^, along with a predicted average resection index of 0.83 ± 0.12. Subsequently, through early post-surgical MRI assessment, the actual mean residual volume was determined to be 0.19 ± 0.05 cm^3^. Notably, the achieved resectability index was remarkably high, measuring 0.99 ± 0.03.

### Survival Analysis

From the 28 patients included in this analysis, a median OS of 532 (39) days [17.7 months], and a median PFS of 300 (40) days [10 months] were achieved. Age showed a positive and statistically significant effect on OS, with a hazard ratio (HR) of 1.07 (95% CI [1.00, 1.14], p = 0.05), indicating that older patients had a slightly higher risk of mortality. Preoperative KPS had a negative and significant impact on survival, with an HR of 0.92 (95% CI [0.87, 0.97], p = 0.00), suggesting that patients with lower preoperative KPS scores had a higher risk of mortality. The only predictor significantly associated with PFS was the preoperative KPS. Patients with lower preoperative KPS scores had a higher risk of progression (HR = 0.94, 95% CI [0.89, 0.99], p = 0.01).

## DISCUSSION

In the present study, our objective was to conduct a meticulous evaluation of the efficacy of nn-2D-IOUS in glioma surgery, specifically examining its influence on the EOR and patient outcomes. Positioned within the context of rapidly advancing neurosurgical technologies, our findings robustly uphold the merit of this traditional modality, asserting that it produces clinical outcomes that are commensurate with those generated by more advanced technologies.

The quality of images procured by the nn-2D-IOUS was deemed satisfactory in most scenarios, with a success rate of 79%. Nevertheless, the delineation of tumor boundaries posed a considerable challenge, yielding successful demarcation in a mere 33% of instances. Despite this limitation, this modality exhibited proficiency in detecting residual tumor tissue in 27% of the cases. In the instances where the technology failed to detect residual tumor, the mean volume of the overlooked residual tumor was strikingly minimal, averaging at 0.29 cm^3^. This data accentuates the promising role of nn-2D-IOUS in surgical contexts, aiding in the distinction between tumor and healthy tissue, and thereby enhancing the potential for safe, maximal resection.

When evaluating the EOR, our findings indicated successful attainment of GTR in a notable fraction (69%) of gliomas, in which surgical intervention was intended for total removal. This included both high and low-grade gliomas, which were non-selected and operated on consecutively, and our adherence to a stringent definition of GTR was consistently maintained. In the specific subgroup of glioblastomas, we introduce an objective measure of surgical difficulty, represented as a tumor resectability index. The resection grades observed within this patient subgroup exceeded anticipated probabilities, thereby substantiating the use of nn-2D-IOUS as a valuable intraoperative tool.

When comparing our results with the existing literature, several comparative points emerge. Solheim et al. ^17^, applied a navigated ultrasound system to an unselected series of 156 high-grade gliomas, yielding an average GTR rate of 37% for the entire cohort, and 63% for the subgroup of gliomas amenable to total removal. Employing 3D navigated IOUS, Bo et al.^18^ reported a series of 47 cases of low-grade gliomas, achieving a GTR rate of 30%.

In a study conducted by Moiraghi et al.^19^, an analysis was carried out on a series of 60 high-grade glioma cases. The use of navigated 2D IOUS was compared against standard neuronavigation without IOUS across 31 and 29 cases respectively. The GTR rate for the group using navigated ultrasound was 61.2%, in contrast to the 44.8% achieved with navigation alone. The study further asserted that navigated ultrasound proved beneficial in detecting residual tumors exceeding 1 cm^3^ in volume.

In a study conducted by Moiyadi et al.^3^, navigated 3D ultrasound was employed across a cohort of 111 patients, encompassing not only high and low-grade gliomas, but also metastases, meningiomas, and other conditions. The overall GTR rate for the cohort stood at 53%, with a higher rate of 75% recorded for the group scheduled for radical surgery. The diagnostic accuracy for the detection of residual tumors was found to be 82.5%, a figure closely aligned with the results from our series, which documented an accuracy of 87%.

With respect to the integration of IOUS with other operative adjuncts, it is worth mentioning the study published by Della Pepa et al^20^. In this study, the author carried out a retrospective comparison of the use of ultrasound with Contrast-Enhanced Ultrasound (CEUS), 5-Aminolevulinic Acid (5-ALA) used individually, and their combination against unassisted microsurgery. They compared these methods in terms of their resection rates in glioblastomas. The GTR rates, when utilizing IOUS combined with CEUS or 5-ALA individually, stood at a mere 11.6%. However, when these tools were combined, the rate escalated to 69.8%, showcasing the synergistic potential of these two techniques. Despite this marked increase, it still fell short of the GTR rate recorded in our glioblastoma series, which was 70% for the entire subgroup and 81% for surgeries aimed at total removal.

In terms of the capacity of intraoperative ultrasound (IOUS) to detect residual tumor tissue, our data indicates a high specificity of 100% along with a commendable sensitivity rate of 68%, as compared with early postoperative MRI results.

A meta-analysis performed by Trevisi et al.^21^, incorporated thirteen studies focusing on both high-grade and low-grade gliomas. The consolidated sensitivity was reported at 72.2%, with a specificity of 93.5%. Among these studies, five utilized non-navigated 2D ultrasound, and no substantial disparities were observed when juxtaposed with the performance of 3D and navigated ultrasound. Munkvold et al.^5^ reported a lower sensitivity rate of 46%, albeit with a reasonable specificity of 85%. De Quintana-Schmidt et al.^22^, in a series of 100 patients employing navigated ultrasound, reported a sensitivity of 94% and a specificity of 100% in detecting residual tumors. It should be noted that their series included various tumor types alongside gliomas.

The integration of navigation into intraoperative ultrasound (IOUS) undoubtedly provides substantial benefits in terms of anatomical orientation. However, the pivotal question remains: does the inclusion of navigation yield significantly different results compared to the achievements made possible by a proficient learning curve utilizing conventional 2D ultrasound?

To date, only one study published by Renovanz et al.^23^ examines the influence of integrating navigation into IOUS on the extent of resection (EOR) in high-grade gliomas. The study encompassed 92 patients, including 32 reoperations. Navigated ultrasound was used in 49 cases, while non-navigated ultrasound was deployed in 44 cases. The authors defined gross total resection (GTR) as resection exceeding 95%, a definition differing from the most recent published guidelines ^12,13^ that we have employed in our current study. The GTR rate for the navigated ultrasound group was 49%, compared to 44% for the non-navigated group, with no statistically significant differences noted.

A study by Miller et al., published in 2007, compared the use of integrated navigation technology with IOUS against non-navigated IOUS in a series of 29 patients with various tumor types, including gliomas, metastases, meningiomas, among others. The conclusion was predominantly subjective, stating that navigation aids in anatomical orientation.

In a randomized controlled trial published by Incekara et al.^24^, the use of 2D IOUS in B-mode (23 patients), whether navigated or not, was compared against another group utilizing only conventional neuronavigation (24 patients) for radical glioblastoma surgery. They reported a GTR rate of 35% in the ultrasound group compared to 8% in the conventional navigation group. A cost-effectiveness analysis published by Eljamel et al.^25^ reported a GTR rate of 73.4% using IOUS compared to 70% with intraoperative MRI, with no significant differences. However, the additional cost per Quality-Adjusted Life Year (QALY) was $665 for ultrasound and $32,955 for intraoperative MRI. Mosteiro et al.^26^ reported similar results, with GTR rates of 70% using intraoperative MRI and 60% for non-navigational intraoperative ultrasound. It should be noted that GTR was defined as a minimum of 90% resection.

Our study reasserts the essential role of technologies such as 2D IOUS in glioma surgery. Although the demand for advanced intraoperative imaging technologies is indeed high, our findings indicate that the proficiency in deploying fundamental tools like IOUS can yield superior patient outcomes.

The limitations of nn-2D-IOUS must not be overlooked. The effectiveness of the technology is largely contingent upon the surgeon’s skill and experience, potentially leading to varying outcomes. As evidenced in our study, the variability in the quality of the images procured and the ability to accurately delineate tumor borders underscore these limitations. Furthermore, the steep learning curve associated with IOUS should not be underestimated. However, these constraints do not necessarily discredit the usefulness of IOUS; instead, they emphasize the importance of comprehensive training and ample experience to optimize its application.

There are a few limitations to consider in our study. Firstly, the retrospective nature of the study introduces potential biases and limitations in data collection. Additionally, the sample size, especially for low-grade gliomas, was relatively small, which may limit the generalizability of our findings. It’s important to note that our study was conducted at a single center, so the results may not be fully representative of other institutions or patient populations. Furthermore, we did not directly compare our intraoperative imaging modality with other available techniques, which could provide further insights. While these limitations should be considered, they do not diminish the value of our findings in providing worthwhile insights into the topic.

Indeed, the relentless advancement of medical technologies plays an essential role in enhancing surgical outcomes in fields as complex as neuro-oncology. Affluent institutions undoubtedly benefit from these innovations. However, our study highlights those traditional, well-established modalities such as 2D-nn-IOUS, when applied skillfully, can yield comparable, if not superior, results. Our findings suggest that a focus on refining and maximizing the use of existing technology can be a practical and effective strategy, especially for resource-constrained settings.

## CONCLUSION

Our research distinctly accentuates the merit and effectiveness of the two-dimensional, non-navigated intraoperative ultrasound within the context of glioma surgery. Notwithstanding the increasing tendency towards more sophisticated imaging technologies, the importance and practicality of nn-2D-IOUS persist to be robust within the domain of neurosurgical oncology. The drive towards acquiring advanced, high-tech instruments should not overshadow the pragmatic benefits and cost-efficiency offered by conventional methodologies.

## Data Availability

A portion of the MRI scans used in this study will be made publicly available through The Cancer Imaging Archive at https://www.cancerimagingarchive.net/

https://doi.org/10.7937/4545-c905

